# Multidimensional Modeling to Maximize Adaptations to eXercise: The M^3^AX Trial Rationale and Study Design

**DOI:** 10.1101/2025.05.27.25328425

**Authors:** Zachary A. Graham, Matthew P. Bubak, Christiana J. Raymond-Pope, Gary R. Cutter, Jeremy S. McAdam, S. Craig Tuggle, Jacob A. Siedlik, Luis G.O. de Sousa, Edward J. Chappe, Kana Meece, Amrit Kaur, Benny S.R. Ruiz, Sophia C. Bamman, Katherine M. Vanselow, Trevor W. Perry, Jorge S. Acosta-Arreguin, Natalie J. Bohmke, Greg J. Addison, J. Michelle Bowers, Rachel L. Wright, Lena D. Fuentes, Jennifer E. Smith, Karyn A. Esser, Benjamin F. Miller, Sue C. Bodine, Marcas M. Bamman

## Abstract

Age-related functional declines are thought to be caused by hallmark biological processes that manifest in physical, mental, and metabolic impairments compromising intrinsic capacity, healthspan and quality-of-life. Exercise is a multipotent treatment with promise to mitigate most aging hallmarks, but there is substantial variability in individual exercise responsiveness. This inter-individual response heterogeneity (IRH) was first extensively interrogated by Bouchard and colleagues in the context of endurance training. Our group has interrogated IRH in response to resistance training and combined training, and we have conducted trials in older adults examining dose titration and adjuvant treatments in attempts to boost response rates. Despite the work of many groups, the mechanisms underpinning IRH and effective mitigation strategies largely remain elusive. The National Institute on Aging (NIA) hosted a focused workshop in 2022 titled “Understanding heterogeneity of responses to, and optimizing clinical efficacy of, exercise training in old adults”. This workshop spurred a dedicated NIA request for applications (RFA) with the major goal “to better understand factors underlying response variability to exercise training in older adults.” We developed a two-phase Sequential Multiple Assignment Randomized Trial (SMART) in response to the RFA that will allow us to classify individual responsiveness to combined endurance and resistance training and interrogate potential mechanistic underpinnings (Phase I), followed by an approach to boost responsiveness (Phase II). Using deep in vivo, ex vivo, and molecular phenotyping, we will establish multidimensional biocircuitry of responsiveness and build predictive models, providing a basis for personalized exercise prescriptions.

## Rationale

Aging is a complex, multifactorial process that increases the risk of developing morbidities and overall mortality^1^. US Census Bureau data indicate from 2010 to 2020 the population of individuals 65+ years of age has grown^2^ and currently makes up 17% of the overall US population (∼56 million), primarily driven by an increase in the number of older adults aged 65 – 74 years. Cardiorespiratory fitness (CRF, VO_2_peak) is a predictor of cardiometabolic disease risk and all-cause mortality with no apparent upper limit of benefit at higher CRF values^3–5^. With aging, there is a gradual decline in CRF which at a certain threshold (18 ml/kg/min) can cross into disability^6,7^. Further, losses of strength and muscle mass that occur with aging (i.e., sarcopenia) are linked to a higher risk of morbidity, disability, and mortality^8–11^. Low CRF and low functional muscle quality (fMQ; strength/muscle mass) are multi-system manifestations of the deterioration of cellular aging hallmarks. Interventions that successfully improve CRF and fMQ could extend healthspan, decrease functional disability and have major health, quality of life, and economic impacts^12^.

The health benefits of exercise training are multipotent, affecting virtually all cells, tissues, and organ systems, as previously reviewed in detail^13^. However, the degree to which individuals positively adapt to exercise can vary, which has been termed inter-individual response heterogeneity (IRH). Our group has demonstrated IRH even during supervised and rigorously conducted laboratory-based progressive exercise training programs in young^14^ and aged individuals^15^. IRH stems from modifiable (e.g., exercise, sleep) and non-modifiable (e.g., age, genetics) factors that drive changes associated with the hallmarks central to aging such as proteostasis and mitochondrial energetics, among others. While exercise training mitigates many of the aging hallmarks, individual differences likely contribute to IRH among older adults. For example, a lack of mitochondrial energetic resources to maintain proteostasis during exercise training may contribute to poor responder status. Aging also disrupts circadian clocks, leading to inflammation and impaired musculoskeletal and metabolic health; however, exercise shows promise for resynchronization^16–18^. Thus, the magnitude of exercise-induced improvements in clock phase synchrony or amplitude may be linked to responder status.

Despite similarly high amounts of effort among individuals during exercise training, individual gains to counter age-related declines in CRF and fMQ vary widely. A better understanding of the features influencing IRH should enable exercise prescription tailored to the individual including personalized rehabilitation. Previous and active large-scale exercise trials studying IRH (e.g., **HE**alth, **RI**sk factors, exercise **T**raining **A**nd **GE**netics, HERITAGE; **Mo**lecular **Tr**ansducers of **P**hysical **A**ctivity **C**onsortium, MoTrPAC) have implemented only endurance training (ET) or resistance training (RT). The HERITAGE study provided a key understanding that there are genetic limiters in an individual’s response to ET^19^. As data emerge from the MoTrPAC trial of over 1500 participants across the adult age spectrum, IRH to ET and to RT is fully expected. The U.S. Department of Health and Human Services (HHS) 2018 guidelines for weekly physical activity urge combined ET and RT, which target CRF and fMQ, respectively, to maximize exercise-induced health benefits^20^. We recently studied IRH in response to combined ET and RT in young adults^14^. However, the impact of combined ET and RT and its effects on IRH has yet to be studied in older adults.

Physiological systems require a sufficient stimulus to induce adaptation. It is possible that IRH partially stems from insufficient exercise volume and/or intensity, suggesting that modifying the exercise stimulus could mitigate poor responsiveness. Prior literature has shown that increasing volume or intensity during ET or RT improves fMQ and CRF, respectively^15,21–24^. For example, in post-menopausal women who underwent six months of ET, increasing the training volume in non-responders resulted in an increase in VO_2_peak, thus eliminating the non-responsiveness to exercise^22^. Among older adults, increasing training volume during RT converts some low-volume non-responders to responders based on MRI-assessed gains in muscle size^21^. Others have also shown greater gains in strength and muscle fiber cross-sectional area (CSA; i.e., hypertrophy) in response to higher RT volume^21^. However, it is unknown if increasing exercise stimuli in a combined ET and RT training program would boost responsiveness among older adults across one or both primary outcomes (CRF and fMQ).

A workshop organized by the National Institute on Aging (NIA) led to a white paper^25^ summarizing the potential underlying and intersecting multi-domain mechanisms that affect IRH. Subsequently RFA-AG-24-045, titled “Elucidating variability of physiologic and functional responses to exercise training older adults,” was released. In response to this RFA, we designed a two-phase Sequential Multiple Assignment Randomized Trial (SMART) to provide evidence-based answers for two primary questions: 1) Phase 1 (Interrogation): What are some of the key mechanisms that drive IRH to exercise in generally healthy older adults?; and 2) Phase 2 (Mitigation): Can easily-administered strategies (e.g., increased exercise intensity/volume, guidance on lifestyle behaviors, stress management) be used to address poor exercise responsiveness? We hypothesize that factors central to aging itself are chief contributors to the multidimensional circuitry that determines whether an individual achieves the minimum clinically important difference (MCID) in CRF and/or fMQ with exercise training. We further hypothesize that altered exercise dosing coupled with lifestyle recommendations will promote attainment of MCIDs among older adults who do not initially respond. To determine the chief contributors to IRH, we will use multidimensional modeling by integrating: targeted cellular/molecular outcomes; inter-tissue communication (extracellular vesicle transcriptomics); epigenomics; free-living behavior [activity, sleep, diet, continuous glucose monitoring, heart rate (HR)/heart rate variability (HRV)]; and clinical outcomes. Individuals who do not attain one or both MCIDs in the 12-week Phase 1 will undergo an additional 10 weeks of training with augmentation via altered exercise dosing and free-living recommendations. We anticipate advances in understanding mechanisms impacting IRH among older adults by: (i) determining the role of aging hallmarks; (ii) defining multidimensional circuitry of influential features; (iii) determining the efficacy of augmentation strategies; and (iv) assessing sustainability of clinically potent CRF and fMQ during 10 weeks of free living.

## Study Overview

This trial is a collaborative effort across three institutions. The Florida Institute for Human and Machine Cognition (IHMC) and the Oklahoma Medical Research Foundation (OMRF) serve as the clinical sites, with the University of Florida (UF) performing *in vitro* primary cell analyses. The overall trial design centers around the ability to classify both CRF and fMQ responder status based on the general proportions from our recent combined exercise dosing trial **(Figure 1)**^14^. We aim to enroll n = 250 sex-matched older adults (60+ yr) to complete 12 weeks of supervised exercise training in Phase 1 followed by an additional 10 weeks of supervised training in Phase II for all who do not attain both CRF and fMQ MCIDs in Phase I. Those who are classified as responders in Phase I (i.e., attain both MCIDs) will be randomized in Phase II to continued supervised training or free-living. Enrollment will be stratified by sex and age group to attain equal sex distribution [50 ± 5% F and M] and at least 30% ≥ 70 yr at each site. The participants will be untrained, defined as no regular exercise training ≥ 2 times/wk in the past year. This trial is registered at clinicaltrials.gov under NCT06507189. The trial design is described in detail below and summarized in **Figure 2**.

**Fig. 1.**
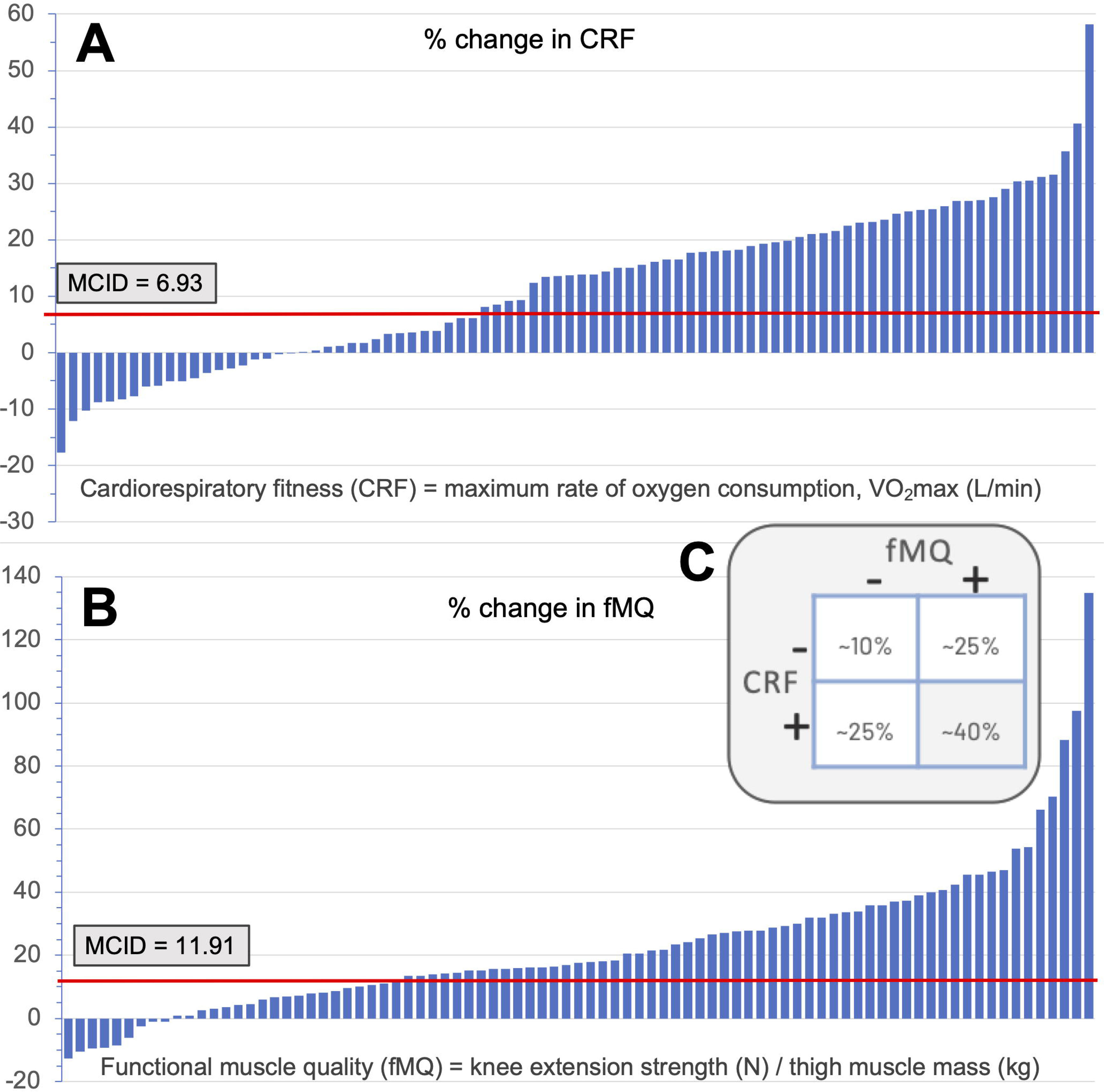
Using the traditional clinical and rehabilitative practice of calculating MICDs to track progress or define treatment/intervention efficacy, we leveraged data from our previous large-scale combined ET+RT clinical trial in young individuals to generate MCIDs of exercise responsiveness. We selected an established distribution-based method to generate MCIDs based on % change from pre- to post-12 weeks of training for A) cardiorespiratory fitness (CRF) and B) functional muscle quality (fMQ). These MCID criteria generated the expected responder breakdown (‘-‘ = did not meet MCID, ‘+’ met MCID shown in the Punnet Square shown in C).

**Fig. 2.**
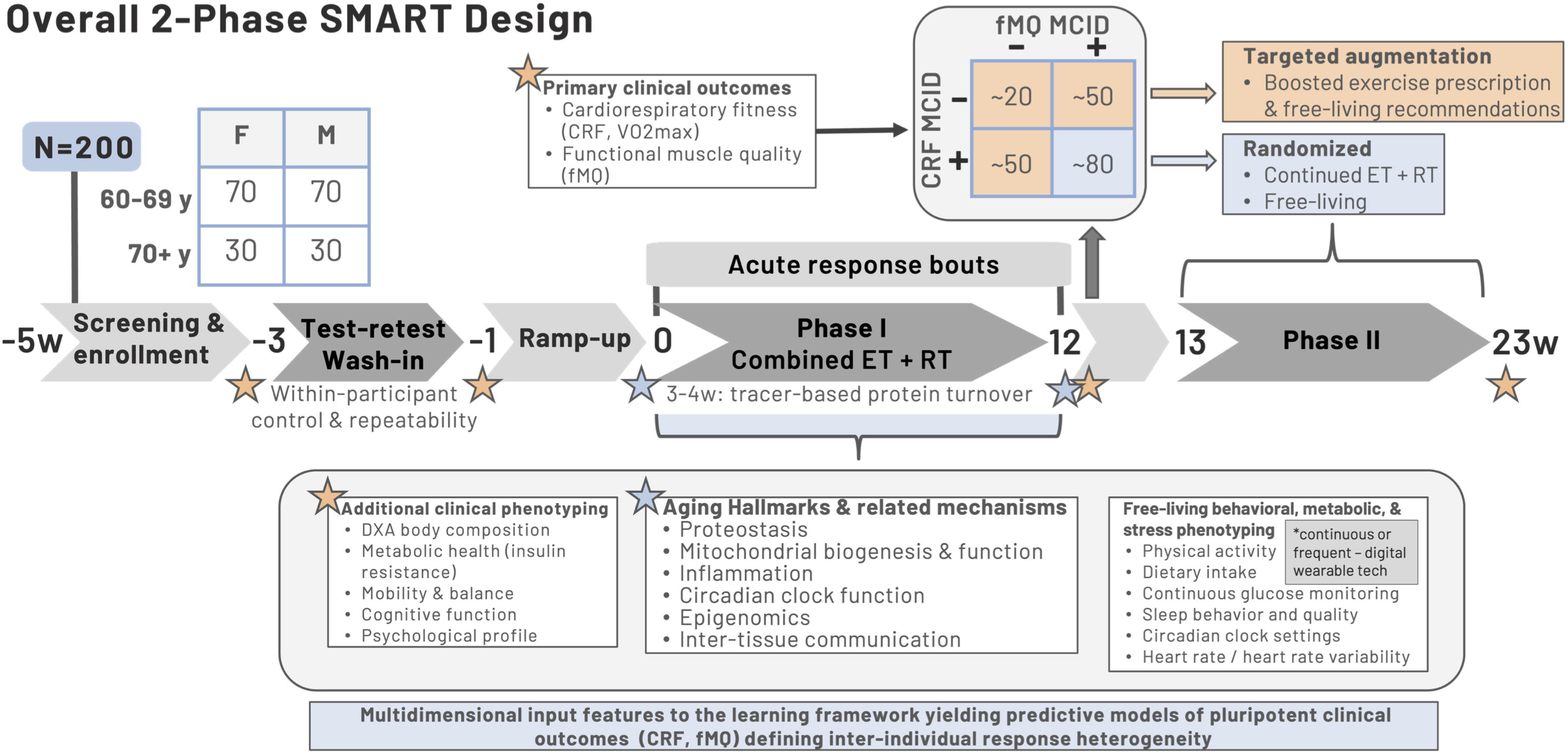
Overall trial design. The 2-phase SMART design is intended to determine what factors and mechanisms underlie inter-individual exercise response heterogeneity (IRH) among older adults. We employ a rigorous exercise prescription of combined endurance and resistance training to ensure direct translatability to public health recommendations. Phase I (12 week) is focused on ***interrogation*** which involves testing targeted hypotheses anchored to hallmarks of aging complemented by a multidimensional learning framework to decipher complex interrelationships among clinical, molecular, and behavioral phenotyping. Our modeling framework will produce integrated, multidimensional circuits predictive of responsiveness that contain modifiable factors for future study and precision exercise medicine. Phase II (10 week) will attempt ***mitigation*** by a targeted augmentation strategy for the older adults who do not attain minimum clinically important difference (MCID) scores for one or both primary outcomes (CRF and fMQ). Those in Phase II who meet both MCIDs will be randomized to test sustainability of training adaptations during free-living (vs. continuing the supervised intervention). Planned enrollment is therefore N=250 to account for up to 20% data missingness (data loss, QA/QC, attrition).

### Inclusion and exclusion criteria

Our aim is to enroll otherwise healthy individuals meeting the following criteria:

#### Inclusion

- Male or female aged 60 yr or above
- Free of chronic disease
- No structured exercise program (2 or more bouts/wk) within the previous 12 months
- Cognitively capable of providing informed consent

#### Exclusion

- Neuromuscular or musculoskeletal disorder that would limit ability to perform the exercise and/or testing bouts
- Cardiopulmonary disorders
- Metabolic diseases including markers of liver disease (ALT > 52 U/l) and type 2 diabetes (HbA1C ≥ 6.5, fasting blood glucose ≥ 126 mg/dl)
- Any other disease or disorder that would influence exercise response (e.g., chronic kidney disease, dementia, current cancer diagnosis or within 2 yr remission, cerebrovascular disease)
- Any current infectious disease
- Life expectancy < 1 yr
- Insulin sensitizing/blood glucose lowering agents such as metformin
- Weight loss medication (e.g., GLP-1 receptor agonist) or other medication that influences appetite or metabolism
- High dose statin (≥40 mg/d simvistatin equivalance)
- Lidocaine allergy
- Regular tobacco use and/or vaping
- Excessive alcohol consumption (3 drinks/d or 7 drinks/wk for females; 4 drinks/d or 14 drinks/wk for males)
- BMI ≥ 30.0 kg/m^2^
- Unable to commit to ∼6 months required to complete the study

### Test-retest wash-in

Randomizing volunteers to a no-exercise control group, which would restrict older adults from exercising for 6 months, was not preferred nor practical; thus in lieu of a no-exercise parallel control group, we implemented a test-retest wash-in prior to the Phase I intervention for rigor to provide within-participant control. The test-retest wash-in includes repeat assessments for most outcomes to prevent unintentional drivers of IRH. For example, it is easy to imagine an individual performing worse than their true capacity during their pre-training baseline VO_2_peak or strength tests due to something as simple as acute test anxiety. After several weeks of training the participant is then comfortable with maximal exercise and thus performs up to their capabilities during post-testing, resulting in IRH being unintentionally magnified. This concern is mitigated by the test-retest wash-in across physiologic, body composition, cognitive, and psychologic/mood domains. All retests occur a minimum of one week after the initial test, with the best performance taken as the true measure. For non-performance dependent tests [e.g., whole-body dual x-ray absorptiometry (DXA) scan], the two tests are averaged to account for technical variation vs. effort or biologic variation.

### Phase I: Exercise prescription

All participants undergo 12 weeks of combined ET and RT (**Figure 3**), with each week containing three sessions. We selected this frequency to meet the HHS guidelines for exercise doses in aging populations^20^. The prescription is anchored to variations in intensity each week, ensuring a higher intensity stimulus is implemented for each mode including weekly exposure to high-intensity interval training (HIIT) and twice weekly exposure to near-maximal contractions during RT. Intensity is a key driver of adaptation in both modes as the production and secretion of exerkines, which influence for inter-tissue communication, is also intensity-dependent^26–28^. Progression of volume and intensity occurs during the ramp-up week and into the first week of training. For RT, the ramp-up period increases the number of sets, repetitions, and intensity to limit excessive muscle damage and soreness. Full volume training is achieved by the end of Week 1 and progression thereafter is based on progressive load increases to maintain target intensity. The RT prescription follows our High-Low-High design we demonstrated to be effective for increasing muscle mass and strength in older adults^15,29^. ET consists of 3x/wk training with sessions 1 and 3 being moderate intensity continuous (steady state) cycling, treadmill, or elliptical at 70-75% heart rate reserve (HRR) for 30 min and session 2 being a 20 min HIIT session on a cycle ergometer (1 min on/off; 10 cycles) targeting 85-90% HRR.

**Fig. 3.**
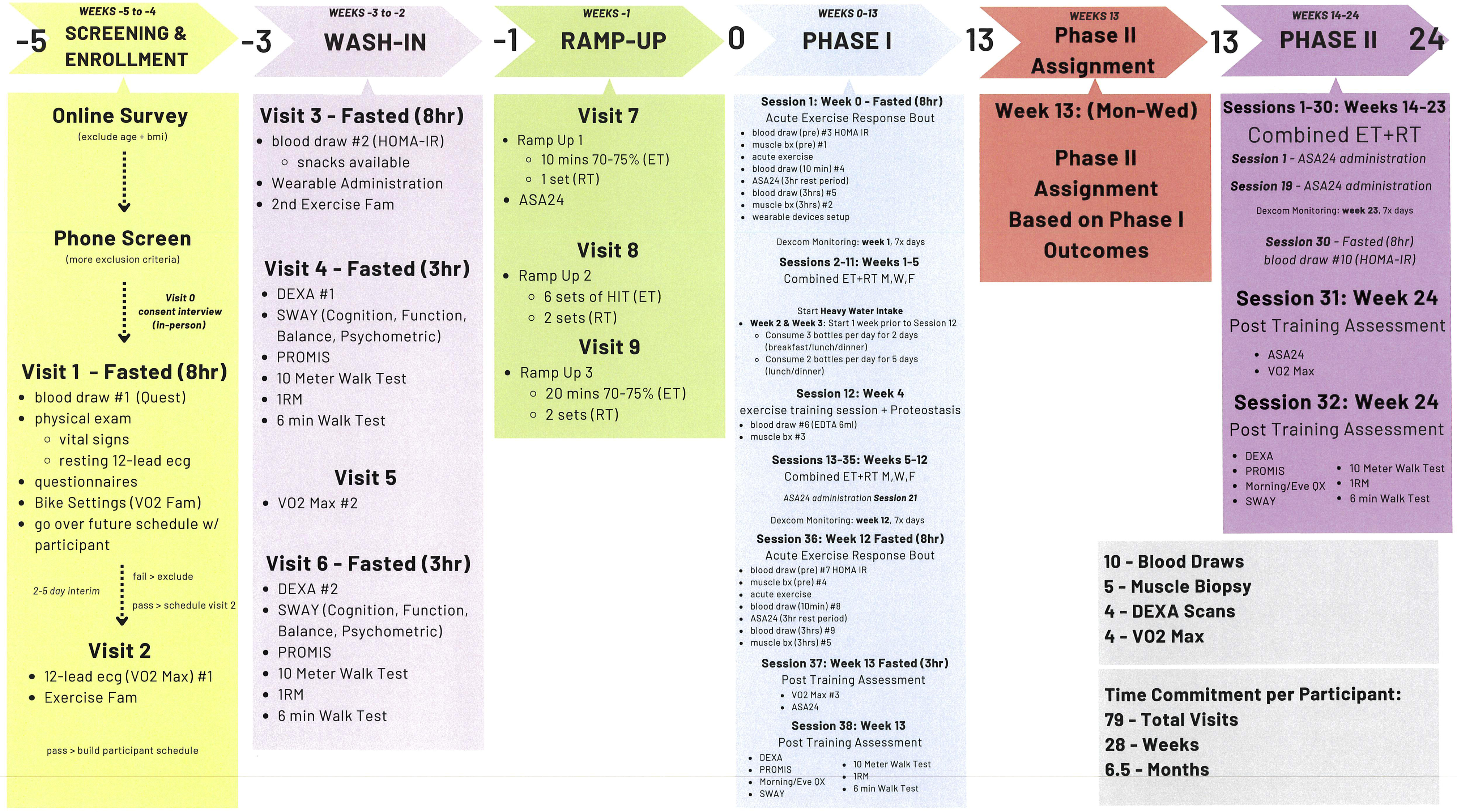
General participant flowthrough and expected burden from screening through end of the study.

**Fig. 4:**
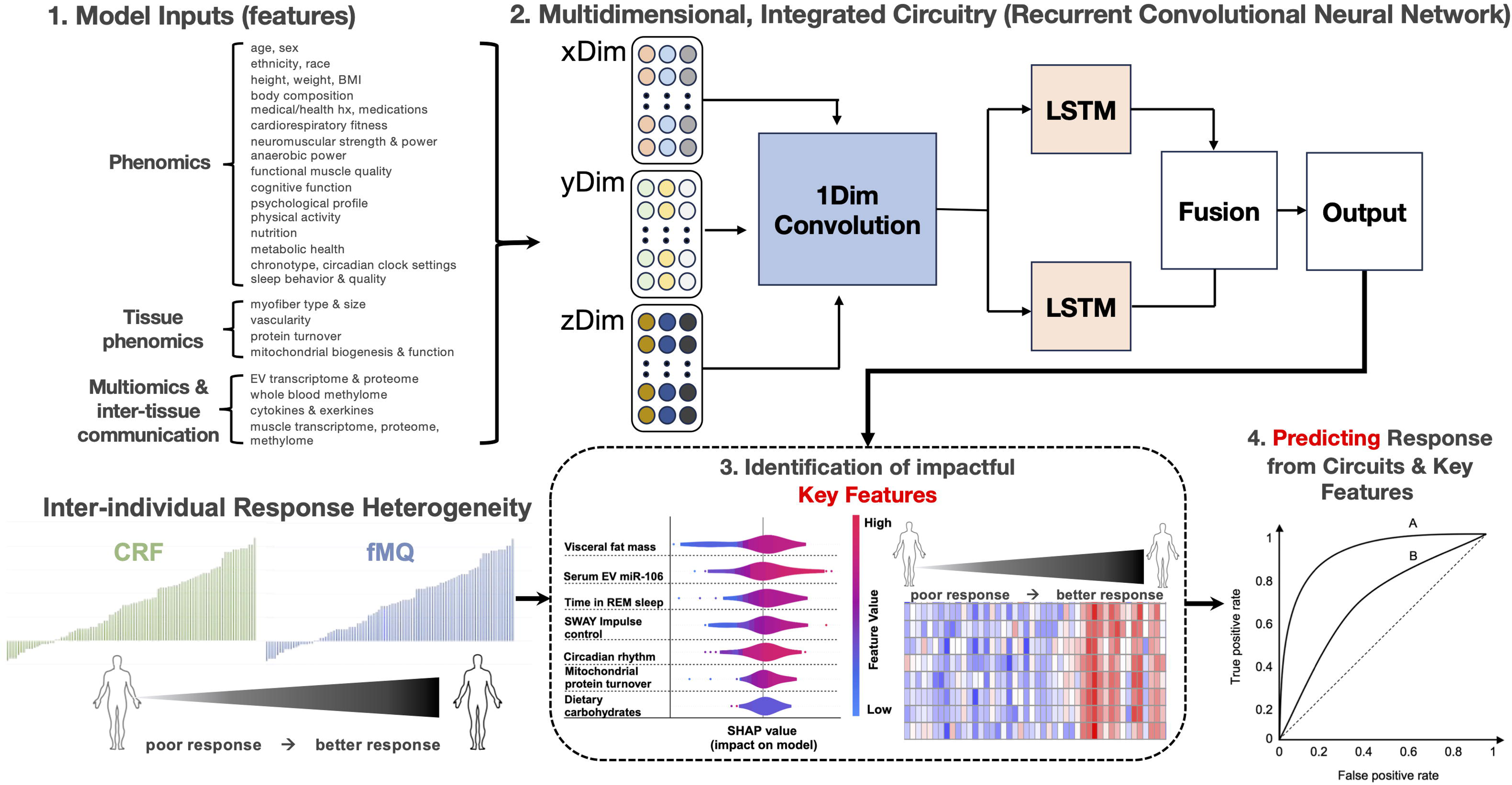
Multidimensional learning framework to model and predict inter-individual exercise response heterogeneity. <u>Step 1:</u> Model inputs are curated independently within each data set, with each variable considered a feature. <u>Step 2:</u> Multidimensional, integrated phenomic, cellular, molecular multiomic and inter-tissue communication circuitry established using Recurrent Convolutional Neural Networks (RCNN). Baseline and longitudinal data from across domains (e.g., physical performance, molecular signatures) are concatenated and passed through a 1D convolutional layer to extract local temporal patterns. The output is then fed into parallel Long Short-Term Memory (LSTM) layers to capture sequential dependencies and temporal dynamics. <u>Step 3:</u> Outputs from each LSTM stream are fused and passed to a final dense output layer for identification of key features as determined using SHapley Additive exPlanations (SHAP) values. <u>Step 4:</u> Selected impactful features are based on two relevant criteria: good fit (i.e., high SHAP values) and actionable/modifiable. The most influential based on strength of relationship with responder classification will serve as inputs for predictive modeling of responsiveness.

Both ET and RT will be progressed per individual by monitoring each session with pragmatic increases in cycling wattage, treadmill speed/grade, and weight lifted. After Phase I, there will be a post-intervention testing week that repeats the battery of pre-training testing to determine responder status by our *a priori* designated MCIDs for CRF and fMQ (6.93% and 11.91% increase, respectively). We expect 50% of the full cohort will only attain one MCID (CRF or fMQ), 40% will attain both, and 10% will attain neither. This predicted distribution is based on our prior trial in young adults^14^.

### Phase II: Mitigation

Each of the four responder classes [(i) CRF^+^/fMQ^+^, (ii) CRF^+^/fMQ^-^, (iii) CRF^-^/fMQ^+^, (iv) CRF^-^/fMQ^-^)] will continue into Phase II, a 10 week program in which the dual CRF^+^/fMQ^+^ responders are randomized to continue supervised training or return to free living and the other three classes continue supervised training with an augmentation to boost the outcome(s) in which they were not responsive (**Figure 3**).

### Dual responder randomization

The ∼40% of participants we expect will meet the MCID criteria for both CRF and fMQ will be randomized following Phase I to either continue progressive, laboratory-based exercise training or return to free living. Those randomized to free living will have no imposed restrictions on activities. As this free-living arm is meant to test sustainability of the intervention, they will be given a single session for recommendations of healthy lifestyle behaviors (described below) and will only return for Phase II post-testing.

### Augmentation

The augmentation strategies for both ET and RT are mostly based on intensity-based approaches with a modest increase in overall volume. We suspect some individuals resistant to gains in maximal performance (on which both CRF and fMQ are based) do not necessarily need more training volume per se, but more exposure to training at a higher intensity to induce demand-based adaptations.

#### ET boosting

Those falling short of the CRF MCID (∼6.93% increase) will have ET intensity boosted during weekly Sessions 1 and 3 only (weekly Session 2 HIIT will remain the same). This will be accomplished by adding 2 x 1 min sprint intervals at 85-90% HRR at the conclusion of the 30 min continuous 70-75% HRR steady state bout. The rationale is to perform non-steady state exercise on a more frequent basis at higher intensities akin to the later stages of a CRF test.

#### RT boosting

Those falling short of the MCID for fMQ (∼11.91% increase) will experience an increase in volume and intensity for the knee extensor exercises during weekly Sessions 1 and 3 while the midweek Session 2 prescription will remain unchanged. Volume will be increased by adding one additional set to the knee extension and leg press exercises. The increase in intensity will be achieved by reducing the target repetition range only in this one added set from 8-12RM to 5-7RM. The rationale here is that the higher contraction intensity will induce near maximal motor unit recruitment during each repetition. The remainder of the RT prescription will remain unchanged.

#### Free-living recommendations

All participants undergoing augmentation will receive lifestyle recommendations to aid in best positioning them for positive training adaptations. These recommendations will focus on reinforcing the importance of sleep quality, a healthy diet with adequate protein intake, monitoring stress levels via wearable device (heart rate HR/HRV), and engaging in free-living physical activity. We will show participants how to interpret and act on their data independently, with formal discussion and interactions with research staff four times during Phase II to keep participants engaged. The CRF^+^/fMQ^+^ participants randomized to free-living will receive the same initial session on lifestyle recommendations and how to interpret and use their data throughout Phase II, but they will not receive further guidance.

We have provided a general per participant visit schedule and testing battery in **Figure 3**.

## Clinical phenotyping

Unless stated otherwise, all clinical phenotyping will be completed at three timepoints: during the test-retest wash-in and at the end of Phases I and II.

### Physical function and performance

CRF and fMQ are indicators of physical function and performance and were selected as primary outcomes due to their established impact on morbidity and mortality as indices of exercise-induced health benefits. They are also directly comparable to outcomes evaluated in MoTrPAC as we use the same cycle ergometer ramp protocols implemented in that trial^30^. fMQ is determined from bilateral 1RM maximum knee extensor strength normalized to bilateral thigh lean mass measured via DXA using our previously published protocols^31–34^. Other performance measures include 1RM strength for the major compound movements, balance using accelerometry with the Sway Medical platform [modified Balance Error Scoring System (mBESS) and the CDC 30 s Chair Stand)], a 10-m maximum gait speed test, and the 6-minute walk test. These direct measurements will be supplemented with NIH PROMIS Short Form (SF) questionnaires related to perceptions of fatigue (SF Fatigue 13a) and physical function (SF Physical Function 8b).

### Metabolic health

Pre-training clinical blood panels [comprehensive metabolic panel (CMP) and complete blood count (CBC) with diff], HbA1c, and HOMA-IR, are used as markers of metabolic health and integrated into our multidimensional modeling (described below). We use DXA for whole-body composition. Additionally, each participant will wear a Dexcom G7 continuous glucose monitor for ∼7 days at the beginning and end of Phase I and at the end of Phase II. Lastly, dietary intake is assessed during the wash-in period and repeated at the beginning, midpoint and end of both Phase I and Phase II using the validated Automated Self-Administered 24-Hour (ASA24) Dietary Assessment Tool (National Cancer Institute, Bethesda, MD).

### Cognitive and psychological/mood profiling

We use the Sway Medical platform to determine changes in basic cognitive function using modules for simple reaction time, impulse control, inspection time, memory, and the Cued Stroop. Sway-based questionnaires related to depression (PHQ-9) and anxiety (GAD-7) are used for psychological and mood profiling. Using NIH PROMIS SF questionnaires, we further profile perceptions of pain (SF Pain Intensity 3a, SF Pain Interference 8a) and sleep quality (SF Sleep Disturbance 8a, SF Sleep Related Impairment 8a). Lastly, we assess circadian phenotyping using the Morningness-Eveningness Questionnaire.

### Systemic inflammation

As one of the aging hallmarks, we will assess systemic inflammation via a 10-plex serum cytokine array at baseline, end of Phase I, and end of Phase II. We will use the MesoScale Diagnostics (MSD) MESO QuickPlex SQ 120MM system and their highly validated V-PLEX panel that allows the simultaneous detection of 10 pro-inflammatory cytokines, including IFN-γ, IL-6, IL-8 and TNF-α, among others, from a single sample using electrochemiluminescence.

### Free-living monitoring

We provide participants with an Oura Ring (Gen4) to wear throughout the trial. The Oura Ring is primarily used to monitor sleep quality (e.g., sleep latency, sleep duration) and stress (i.e., sleep HR and HRV) while also collecting free-living physical activity using walking equivalency (i.e., transforms all detected measurements of physical activity into the energy equivalence of steps), body temperature, respiratory rate and oxygen saturation.

## Cellular and molecular phenotyping

The cellular and molecular phenotyping will be completed from biospecimens collected at the beginning and end of Phase I.

### Mitochondrial function

We will perform high-resolution respirometry with Oroboros O2K respirometers to profile saponin-permeabilized muscle bundle respiration using a series of mitochondrial respiratory flux assays. Protocol 1 will evaluate submaximal and maximal complex I, complex II, maximal coupled oxidative phosphorylation, and uncoupled electron transport system capacities using our previously described protocol^35–37^. We will determine ADP sensitivity using Michaelis-Menten Kinetics to calculate apparent Km and Vmax. Protocol 2 will measure complex I-linked H_2_O_2_ emissions using succinate-induced complex I electron backflow^38^. We will also determine the ability of mitochondria to suppress maximal H_2_O_2_ emissions by 50% using one-phase exponential decay analysis to calculate half maximal inhibitory concentration (IC_50_). Protocol 3 uses a creatine kinase energetic clamp technique as previously described.^39,40^. The creatine kinase clamp leverages the enzymatic activity of creatine kinase to titrate the extra-mitochondrial ATP/ADP ratio to measure mitochondrial flux demands across a range of physiological ATP free energy states, essentially conducting an ex vivo ‘stress test’. Lastly, OMRF will perform an ATP-O respiratory assay to determine the changes in ATP synthesis efficiency across different free energy energies^39,41^. Conducting these assays in parallel allows for the determination of where IRH may be limiting the mitochondrial respiratory control and/or conductance across a range of substrate conditions and ATP free energy states.

### Primary muscle progenitor cell culture

Muscle progenitor cells will be isolated from fresh tissue at IHMC and shipped to UF for circadian rhythm profiling using lentiviral circadian clock luciferase reporter assays. These assays generate bioluminescence emissions that will be collected every 15 min for the length of recording (3-4 d) with analyses targeted at the timing of the cellular clock, amplitude, and day-to-day stability (onset, degree of activation, regularity, respectively)^42,43^ to identify changes in circadian and clock functions in the context of exercise training. Furthermore, we will determine whether the starting state of clock function and/or the ability of the circadian clock adapt to exercise training contribute to IRH.

### Muscle histology and targeted molecular assays

We will determine changes in myofiber type and size, fibrosis, vascularity, senescence/DNA damage, and the presence of resident stem cells and inflammatory cells including M1 macrophages prior to and following Phase I. As muscle inflammation may be a major effector of exercise responsiveness, we will perform targeted analyses of protein and gene expression of TNFα, Fn14, and IL-6 and associated intracellular signaling markers (p-STAT3, p-p65 NF-κB).

### Skeletal muscle proteostasis and proteomics

Dysregulated proteostasis is a hallmark of aging with exercise training extensively remodeling the skeletal muscle proteome^44,45^. We will measure changes in proteostatic mechanisms in a subset of participants (n = 100) using muscle biopsies taken before training and ∼4 weeks into Phase I. The 4-week timepoint was selected since it gives us an opportunity to observe heterogeneity in protein synthesis rates during the adaptive phase of exercise training. Body water enrichment will be determined from blood plasma collected at the time of the biopsy (i.e., 4 weeks into Phase I). We will use 70% deuterium oxide stable isotope tracer as we have previously done in exercise studies in aged individuals^35^. Differential centrifugation of the muscle sample allows for the detection of synthesis rates of myofibrillar, sarcoplasmic, and mitochondrial proteins. In addition, our group has developed targeted proteomic panels to measure quantitative (in µg/mg of tissue) changes in individual proteins as well the turnover of those individual proteins^46–48^. The addition of a protein turnover measurement is an important aspect because protein synthesis and breakdown are two of the primary proteostatic mechanisms. For skeletal muscle, we will use protein panels that focus on mitochondrial adaptations, substrate flux, ribosomal biogenesis, and redox balance, among others.

### Inter-tissue communication and muscle transcriptomics and epigenomics

To assess inter-tissue communication, we will perform plasma extracellular vesicle (EV) long and small RNA-Seq during acute response studies surrounding the week 0 (before training) and week 12 (after training) exercise sessions of Phase I on a subset (n = 100) of participants. These timepoints will have EV profiling before exercise, then immediately and 3 h post-exercise. These acute response studies will also include skeletal muscle total RNA-Seq at pre-exercise and 3 h post-exercise timepoints. Baseline epigenomics of skeletal muscle will be determined via ATAC-Seq at week 0 in the fasted morning pre-exercise biopsy as a major component for our multidimensional modeling. A schedule of all biospecimen collections can be found in **Figure 3** and **Supplemental Table 1**.

## Molecular Multiome

This trial is well-powered to detect differential signatures across the ‘omics platforms and integrated multiome. The R package *limma* will be used to perform linear mixed modeling for normalized and transformed transcriptomics and proteomics data to compare differential expression and abundance over time. Analyses will be performed across a subset of n =100 and all steps will be blocked for participant study ID to control for between-participant variability. Significant features will be based on a fold-change (absolute log_2_fold-change > 1, corresponding to a doubling or halving of protein abundance/or RNA expression) and Benjamini-Hochberg-adjusted FDR < 0.05 criteria.

We will further leverage Pathway-Level Information ExtractoR (PLIER) for pathway-informed dimension reduction and generation of latent variables (LVs) for annotated features from the transcriptome and proteome. We will also complete uninformed singular value decomposition (SVD) analyses of a matrix combining all normalized multiomics platforms with equal relative scaling (e.g. z-scores). The ability of PLIER and SVD-alone to deconvolute data from tens of thousands of features to a few dozen LVs allows us to perform standard mixed-model statistics on LV membership to discern important group * time interaction and timecourse effects. Tukey’s multiple comparison testing will be used for follow-up testing to identify differences from our mixed-model outcomes, our statistical criteria for statistically significant LVs will be a Benjamini-Hochberg-adjusted FDR < 0.05.

## Multidimensional learning framework: Recurrent Convolutional Neural Networks (RCNN)

The overall goal of understanding response heterogeneity in CRF/fMQ through predictive modeling will be conducted on binary classification (responder vs. non-responder) and continuous change (e.g., ΔCRF, ΔfMQ). We expect that Phase I will yield a distribution of participants meeting MCIDs of: (i) both CRF and fMQ (40%); (ii) CRF only (25%); (iii) fMQ only (25%); or (iv) neither CRF nor fMQ (10%). These will be modeled using phenome only, molecular multiome only, and then following complete integration using a recurrent convolutional neural network (RCNN) to capture both temporal dynamics and cross-modal interactions.

RCNNs are uniquely suited for this task as they combine the feature extraction strength of convolutional neural networks (CNNs) with the temporal modeling capabilities of recurrent neural networks (RNNs). This allows for simultaneous inclusion of variables with differing sampling frequencies such as static features (e.g., baseline molecular signatures), low frequency repeated measures features (e.g., 1RM) and high frequency measures (e.g., sleep). We will organize these features into parallel subnetworks in the RCNN and unify them in a fusion layer to produce final predictions. We will utilize long-short term memory (LTSM) to address vanishing and exploding gradients that are known to be challenges when implementing RNN on high dimensional data. Hyperparameters (epochs, learning rate, number layers, number units, batch size, dropout rate) will be tuned using cross validation to determine optimal parameter sets.

To address imbalanced sample sizes and non-uniform temporal resolution between datasets, we will incorporate transfer learning and domain adaptation techniques, as previously demonstrated to be effective in similar biomedical applications^49–51^. This architecture not only improves predictive performance but also enhances biological interpretability by identifying latent features that drive inter-individual variability in training response. Ultimately, this model is designed to learn response heterogeneity from multiple data dimensions, including: (i) baseline clinical phenotypes; (ii) longitudinal physiological adaptations; (iii) time-series molecular signatures during acute exercise; and (iv) resting-state molecular adaptations post-intervention.

### Data Preprocessing and Input Construction

Each data modality will undergo preprocessing pipelines appropriate to its structure. Phenomic data will be standardized and imputed for missingness. Molecular multiomic data will undergo normalization (e.g., log2 CPM, TPM, or z-score), filtering for low variance features, and batch effect correction using empirical Bayes methods when necessary.

### Evaluation and Interpretability

Model performance will be evaluated using stratified k-fold cross-validation. For binary classification, we will report area under the ROC curve (AUC), accuracy, precision, recall, and F1 score. For continuous outcomes, root mean squared error (RMSE) and R-squared (*R*²) will be calculated. Model interpretability will be assessed using feature importance derived SHapley Additive exPlanations (SHAP) values. This RCNN-based integrative modeling pipeline aims to balance predictive accuracy with interpretability, allowing both discovery of novel biological mechanisms and practical identification of features relevant to intervention response.

## Conclusions

The M^3^AX Trial is an innovative approach to leverage rigorous and highly supervised combined multi-modal exercise training with deep phenotyping for modifiable factor identification to improve exercise responsiveness in aged individuals. Supported by machine learning and predictive modeling, we expect to find novel relationships within and across domains that will provide translatable information to prospectively apply at an individual level in future exercise prescriptions.

## Data Availability

This clinical trial protocol paper contains no novel data.

## Acknowledgments

This work was supported by the National Institutes of Health (1R01AG089192) to M.M.B., B.F.M., S.C.B. M.M.B is the Contact PI of the trial and will oversee the overall project as well as the team at IHMC. B.F.M and S.C.B are MPIs for the trial and will oversee the project at OMRF. K.A.E is the sub-award PI for the UF site. G.R.C. is the clinical trialist. G.J.A and J.E.S. are the trial clinicians. S.C.T. is the IHMC program manager and C.J.R-P is the OMRF program manager. E.J.C., B.S.R.R, and L.G.D.S are the lead trial interventionists. R.L.W., J.M.B, and L.D.F. are the trial coordinators. Z.A.G., M.P.B., C.J.R-P, N.J.B. K.M., A.K, K.M.V. and S.M.B. will conduct wetlab experiments. J.S.S. is the computational biologist. J.S.M., J.S.AA and T.W.P will oversee data management. J.S.M., J.S.S., and G.R.C. will perform statistical analyses.

Z.A.G, M.P.B., and C.J.R-P. prepared the manuscript. M.M.B, B.F.M., S.C.B., G.R.C and K.A.E. edited/revised manuscript. All authors approved the final version.

