## supplemental table 1 for "Multidimensional Modeling to Maximize Adaptations to eXercise: The M^3^AX Trial Rationale and Study Design"

**Supplemental Table 1. Biospecimen processing and allocation**

| **Time Point** | **Clinical Site** | **Biospecimen ID** | **Biospecimen Type** | **Laboratory**  **Assay(s)** |
| --- | --- | --- | --- | --- |
| Screening Visit 1 | IHMC and OMRF | **BD1** | Blood | CBC, CMP, and Insulin |
| Test-Retest  Visit 4 | IHMC and OMRF | **BD2** | Blood | Glucose (SST) and Insulin (SST) |
| Phase 1 Visit 1 (Pre) | IHMC and OMRF | **BD3** | Blood | Glucose (SST), Insulin (SST), 1-4ml EDTA, 2- 6ml EDTA, 1-10ml SST, 1-8ml CPT |
| Phase 1 Visit 1 (Pre) | IHMC and OMRF | **Bx1** | Muscle | 1 x 15 mg for high resolution respirometry  1 x 20 mg for proteostasis  1 x 20-30 mg long and small RNA-Seq / RT-PCR,  1 x 20-30 mg CNIA  ≥1 x 30 mg biorepository  1 x 30-50 mg mount for IHC |
| Phase I Visit 1 (10 min Post) | IHMC and OMRF | **BD4** | Blood | 1-4ml EDTA, 2- 6ml EDTA, 1-10ml SST, 1-8ml CPT |
| Phase I Visit 1  (3 Hour Post) | IHMC and OMRF | **BD5** | Blood | 1-4ml EDTA, 2- 6ml EDTA, 1-10ml SST, 1-8ml CPT |
| Phase I Visit 1  (3 Hour Post) | IHMC | **Bx2** | Muscle | 1 x 20-30 mg long and small RNA-Seq / RT-PCR,  1 x 20-30 mg CNIA  ≥1 x 30 mg biorepository |
| Phase 1 Visit 12 | IHMC and OMRF | **BD6** | Blood | Heavy Water |
| Phase 1 Visit 12 | IHMC and OMRF | **Bx3** | Muscle | Proteostasis |
| Phase 1 Visit 36 (Pre) | IHMC and OMRF | **BD7** | Blood | Glucose (7ml SST), Insulin (7ml SST), 1-4ml EDTA, 2- 6ml EDTA, 1-10ml SST, 1-8ml CPT |
| Phase 1 Visit 36 (Pre) | IHMC | **Bx4** | Muscle | 1 x 20-30 mg long and small RNA-Seq / RT-PCR  1 x 20-30 mg CNIA  ≥1 x 30 mg biorepository  1 x 30-50 mg mount for IHC |
| Phase 1 Visit 36 (10 min Post) | IHMC and OMRF | **BD8** | Blood | 1-4ml EDTA, 2- 6ml EDTA, 1-10ml SST, 1-8ml CPT |
| Phase 1 Visit 36 (3 Hour Post) | IHMC and OMRF | **BD9** | Blood | 1-4ml EDTA, 2- 6ml EDTA, 1-10ml SST, 1-8ml CPT |
| Phase 1 Visit 36 (3 Hour Post) | IHMC | **Bx5** | Muscle | 1 x 20-30 mg long and small RNA-Seq / RT-PCR  1 x 20-30 mg CNIA  ≥1 x 30 mg biorepository |
| Phase 2 | IHMC and OMRF | **BD10** | Blood | Glucose (SST), Insulin (SST) |

BD: blood draw; Bx: biopsy
